# Acceptability and perceptions of artificial intelligence in organized breast cancer screening: a study of French women

**DOI:** 10.64898/2026.06.07.26354883

**Authors:** Aurélie Jean, Adeline Merceron, Agathe Le Saux, Étienne Mercier, Philippe Benillouche

## Abstract

This study aims to assess women’s perceptions of artificial intelligence (AI) used in breast cancer screening in France by examining their knowledge of AI and the barriers to their participation in organized screening. The results of a survey conducted in June 2025 among a national sample of 2000 women (aged 40-75) reveal limited participation and persistent concerns among women. Nevertheless, despite a low awareness of specific AI applications, a large majority of the women surveyed are very favorable to the use of AI in breast cancer diagnosis, even considering it a lever to increase screening participation.

## INTRODUCTION: CONTEXT AND CHALLENGES

In France, 10 million women aged 50 to 74 are covered by the organized screening program that invites them to undergo a mammogram every 2 years. The participation rate in organized breast cancer screening in France is declining (52% in 2010, 48% in 2023, and 44% in 2024) (1), well below the 70% recommended by the European Union. Several factors explain this low participation (2): the difficulty of reconciling examinations with geographical and scheduling constraints, the fear of pain related to the mammography examination, the fear of ionizing radiation, the fear of the diagnosis, or a certain skepticism about the effectiveness of screening. This skepticism can be explained, among other things, by the existence of so-called interval cancer, which represents about 20% of cancers diagnosed today (source: INCA 2025), detected between two screening mammograms, and whose possibility - wrongly - reduces the confidence of some women in the effectiveness of organized screening, due to a negativity bias. In addition to the relatively low participation rate in the organized screening program, mammograms are also performed outside this framework. These individual screening practices may be driven by the presence of specific risk factors in some women or by personal preferences and clinical considerations in others, resulting in mammography being conducted independently of the national organized screening program.

In parallel, recent years have witnessed significant advances in artificial intelligence applied to computer-aided diagnosis and individualized risk assessment (3)(4)(5)(6), profoundly reshaping contemporary approaches to medicine (7). In clinical practice, these advanced tools enhance diagnostic and therapeutic processes, enabling more accurate, predictive, and personalized care (8). As such, they open new avenues for preventive medicine, with potential benefits for patients, healthcare professionals, and health systems alike (9).

When properly designed, AI tools can enhance radiologists’ diagnostic confidence by supporting the interpretation of medical images. This increased confidence is also closely linked to risk management considerations, particularly those related to algorithmic bias (10)(11), which may result in technological discrimination in which AI systems perform less effectively for certain population groups, including women, depending on factors such as age or ethnic background. In this context, algorithmic explainability (12) - which refers to the ability to understand and interpret the decision-making logic of AI systems - along with robust governance frameworks, plays a key role. These governance approaches include best practices for pre-deployment testing to ensure algorithmic fairness and guarantee equitable system performance across all individuals (13).

In the past, studies conducted in several countries have examined the perception of patients, regardless of gender, regarding the use of AI in healthcare (14)(15). Generally, these studies highlight patients’ confidence in AI to assist doctors rather than replace them (16). Among the concerns, many patients fear the protection of their medical data, which is highly sensitive personal data under European regulations. To date, no studies have been identified that specifically examine French patients’ perceptions of artificial intelligence (AI), nor any research worldwide focusing on women’s perceptions of AI in the context of breast cancer diagnosis.

## METHODS

### Study Type and Participant Recruitment

This study was conducted in collaboration with Ipsos bva. It was conducted from June 24 to 27, 2025, among a representative national sample of 2,000 women aged 40 to 75 living in metropolitan France. Women aged 40 to 49 were included in the sample to capture the perspectives of the upcoming cohort of women eligible for organized breast cancer screening in France, as well as those who are already undergoing screening based on recommendations from their general practitioner or gynecologist.

The sample was constructed using the quota method based on the following criteria: age, urban area category (*table 1*), socio-professional category (*table 2*), and region (*table 3*).

**Table 1:** Weights used within the sample based on the criteria of age and urban area category. (Insee data, 2021 census)

| Age |  |  |  | Urban area Category |  |  |  |  |
| --- | --- | --- | --- | --- | --- | --- | --- | --- |
| 40-49 years old | 50-59 years old | 60-69 years old | 70-75 years old | Rural areas | 2,000 to 20,000 inhabitants | 20,000 to 100,000 inhabitants | Over 100,000 inhabitants | Parisian agglomeration |
| 29% | 30% | 28% | 13% | 23% | 19% | 14% | 29% | 15% |

**Table 2:** Weights used within the sample based on the criterion of the individual’s socioeconomic status (Insee data, 2021 census)

| Socioeconomic status |  |  |
| --- | --- | --- |
| Lower socioeconomic groups | Higher socioeconomic groups | Inactive |
| 28% | 29% | 43% |

**Table 3:** Weights used within the sample based on the criterion of the region of residence. (Insee data, 2021 census).

| Region of France |  |
| --- | --- |
| Ile-de-France | 17% |
| Centre-Val de Loire | 4% |
| Bourgogne-Franche-Comté | 4% |
| Normandie | 5% |
| Hauts-de-France | 9% |
| Grand Est | 9% |
| Pays de la Loire | 6% |
| Bretagne | 5% |
| Nouvelle-Aquitaine | 10% |
| Occitanie | 10% |
| Auvergne-Rhone-Alpes | 12% |
| Provence-Alpes-Côtes d'Azur / Corse | 9% |

### Data collection

Data was collected via an online survey administered through Ipsos bva’s Online Access Panel, in compliance with the General Data Protection Regulation (GDPR). The questionnaire, made in French (provided upon request), consisting of 24 questions, was developed and validated by the authors. It was then distributed online by the company Ipsos bva, which was responsible for programming the questionnaire, and for data collection and processing. The questionnaire was organized into 3 dimensions: women’s behavior regarding breast cancer screening, women’s knowledge of AI applications in health, and women’s expectations for AI’s contribution to organized breast cancer screening. Since the sample was recruited by Ipsos bva, 100% of participants completed the questionnaire.

### Data analysis

The results were analyzed and presented as a percentage, aggregated from the anonymized database.

## MAIN RESULTS

### Women’s behavior and barriers to screening

- **Participation in organized screening and frequency:** Only 59% of the women surveyed reported undergoing a mammogram at least every 2 years. This figure is consistent with national statistics of 44% participation in organized screening.
- **Level of information on screening:** The majority of women (77%) appear to be well informed about the need to be screened every two years, but 6% think there is no screening frequency.
- **Major obstacles:** The main barriers to more frequent screening are lack of awareness (27% “*don’t think about it*”), apprehension about pain (21%), embarrassment due to modesty (14%), and fear of the result (15%).
- **Communication effectiveness:** France’s national health insurance invitation to screening prompted 74% of women to have a mammogram. Furthermore, 80% of women who are not up to date would get screened if a doctor recommended it, given their age or risk, highlighting the crucial role of the healthcare professional.

### Knowledge and perception of AI in health and screening

- **Limited knowledge of AI:** 73% of women have heard of the use of AI in the field of health, but only 24% know of its precise applications.
- **Positive anticipated effects of AI:** Despite limited knowledge, very strong support is observed: nearly 80% of women believe that AI will have positive effects on early disease detection (79%) and on the accuracy of diagnoses (76%). Furthermore, 76% are in favor of their doctor using AI to analyze their images.
- **AI as a lever for screening participation:** 66% of women see the value of AI in breast cancer detection, and 60% consider AI as an additional argument that could encourage them to get screened.

### Future expectations and fears regarding AI in screening

- **Strong future support for AI in screening:** Nearly 80% of women would be in favor of using AI to analyze their mammograms for earlier cancer detection (78%).
- **Fears and reservations regarding the use of AI in screening:** The main fear concerns the confidentiality of medical data, expressed by 43% of the women surveyed. Also, 45% of women fear misuse of the data collected by AI for non-medical purposes.

## DISCUSSION

The findings of this survey reveal a mixed picture of breast cancer screening in France, characterized by ongoing barriers to participation in the organized screening program and strong potential acceptance of artificial intelligence (AI) as a future facilitator. The reasons reported for low participation in the national screening program are not solely information-related but also involve personal and psychological factors affecting women.

An interesting aspect of this study is the remarkable enthusiasm among women for integrating AI into breast cancer screening. Despite still limited knowledge of its precise applications (only 24%), a very large majority of women anticipate positive effects on early detection (79%) and diagnosis (76%). Also, 60% of women consider AI an incentive to undergo screening, and 80% reported being in favor of using AI to analyze their mammograms and enable earlier detection. This suggests that AI could act as a catalyst to improve screening participation, as it may be perceived by women as a marker of improved accuracy and safety.

Nevertheless, 43% of the women surveyed are concerned about the confidentiality of medical data. It is crucial that health centers ensure the security of the data they share with patients to reassure them. Otherwise, AI could become a barrier, canceling out the intended positive effect of technological innovation.

## CONCLUSION

The results confirm low participation in organized screening, primarily driven by insufficient targeted information and fears about the examination itself. Obviously, strengthening the role of the attending physician in personalized recommendations and the deployment of Awareness campaigns targeting psychological barriers such as pain, embarrassment, and anxiety should be further explored. The integration of artificial intelligence represents a strong lever for improving screening participation rates, provided that patients are adequately reassured. It is crucial to communicate the benefits of AI for diagnostic accuracy and to break myths (such as the replacement of radiologists), while ensuring data confidentiality.

## Data Availability

All data produced in the present study are available upon reasonable request to the authors

## ACKNOWLEDGMENTS AND SOURCES OF FUNDING

The authors thank **La Roche-Posay** for their indispensable financial support in conducting this survey.

## AUTHORS’ CONTRIBUTIONS

Dr. Aurélie Jean (Ph.D.), Adeline Merceron, Agathe Le Saux, Étienne Mercier and Dr. Philippe Benillouche (M.D.) contributed to the development, conduct and analysis of the survey.

